# Prediction of heart transplant rejection from routine pathology slides with self-supervised Deep Learning

**DOI:** 10.1101/2022.09.29.22279995

**Authors:** Tobias Paul Seraphin, Mark Luedde, Christoph Roderburg, Marko van Treeck, Pascal Scheider, Roman D. Buelow, Peter Boor, Sven H. Loosen, Zdenek Provaznik, Daniel Mendelsohn, Filip Berisha, Christina Magnussen, Dirk Westermann, Tom Luedde, Christoph Brochhausen, Samuel Sossalla, Jakob Nikolas Kather

## Abstract

**Background and Aims:** One of the most important complications of heart transplantation is organ rejection, which is diagnosed on endomyocardial biopsies by pathologists. Computer-based systems could assist in the diagnostic process and potentially improve reproducibility. Here, we evaluated the feasibility of using deep learning in predicting the degree of cellular rejection from pathology slides as defined by the International Society for Heart and Lung Transplantation (ISHLT) grading system.

**Methods:** We collected 1079 histopathology slides from 325 patients from three transplant centers in Germany. We trained an attention-based deep neural network to predict rejection in the primary cohort and evaluated its performance using cross validation and by deploying it to three cohorts.

**Results:** For binary prediction (rejection yes/no) the mean Area Under the Receiver Operating Curve (AUROC) was 0.849 in the cross-validated experiment and 0.734, 0.729 and 0.716 in external validation cohorts. For a prediction of the ISHLT grade (0R, 1R, 2/3R), AUROCs were 0.835, 0.633 and 0.905 in the cross-validated experiment and 0.764, 0.597, 0.913, and 0.631, 0.633, 0.682, and 0.722, 0.601, 0.805 in the validation cohorts, respectively. The predictions of the AI model were interpretable by human experts and highlighted plausible morphological patterns.

**Conclusions:** We conclude that artificial intelligence can detect patterns of cellular transplant rejection in routine pathology, even when trained on small cohorts.

## Introduction

In patients with end-stage heart failure, organ transplantation constitutes the desired curative treatment concept [1]. This has been made possible in recent decades, in particular, by the advent of new immunosuppressive drugs, which can ensure long-lasting organ preservation. However, organ rejection by the host immune system remains one of the major complications in these patients [2]. Despite the increasing importance of noninvasive methods in the detection of graft rejection, endomyocardial biopsy remains the gold standard for detecting rejection, especially in the first year after transplantation [3,4]. The pathological assessment of such specimens is reserved for highly specialized pathologists and has massive clinical consequences. In 1990 the International Society for Heart and Lung Transplantation (ISHLT) published a guideline for histopathologic diagnosis of acute cellular rejection to standardize this assessment, which has been revised in 2004 [5]. Nevertheless, the purely subjective assessment of pathological sections has certain disadvantages, such as the dependency on appropriately trained experts, as well as remaining inter- and intra-observer variability [6]. In addition, endomyocardial biopsies are obtained either as routine surveillance protocol diagnostics or as a diagnostic investigation in patients with allograft dysfunction and clinically suspected rejection.

Computer-based image analysis programs can potentially support pathology experts in performing diagnostics. In several histopathological applications, it could be shown that such computer-based image analysis programs can show a high level of concordance with human observers, and in some cases the combination with the human experts can improve the consistency of the findings.[7] In particular, the technology of artificial neural networks has brought very good results in many clinically relevant prediction tasks in recent years [8,9]. A recent extension of this technology is the so-called attention-based multiple instance learning [10], in which the artificial neural network can learn which areas of the whole slide image are more relevant than other areas [11,12].

In contrast to solid tumors, in which many studies have examined computer-based prediction of clinically relevant biomarkers in the last three years [9], there are only comparatively few studies in the context of transplantation medicine. Precedent cases exist in the prediction of organ rejection after kidney transplantation [13], as well as applications of simple, handcrafted feature based image analysis methods to cardiac biopsies after transplantation [14,15]. A recent study by Lipkova et al. used the Deep Learning pipeline “CRANE” to predict cardiac allograft transplantation, yielding a very high and clinical-grade performance [16,17].

However, several open questions remain regarding the data requirements to train such systems, as Lipkova et al. trained their system on thousands of patient samples, but this large number of samples is rarely available. Additional questions remain open regarding the generalizability of such systems, and the biological interpretability which can be drawn from their predictions. Finally, new technical approaches such as self-supervised learning (SSL) to pre-train pathology Deep Learning models could yield an improved performance [18], but this has not yet been evaluated in prediction of cardiac allograft rejection.

In the present study, we collected four cohorts from three hospitals of cardiac transplant patients undergoing cardiac biopsy routinely and based on clinically relevant changes. We trained our own SSL-attention-based Deep Learning pipeline as well as CRANE on these data and evaluated the predictive performance regarding the presence of cellular transplant rejection.

## Materials and Methods

### Patient cohorts and experimental design

In this study we included four case series (“patient cohorts”) from three different medical centers in Germany. The first cohort was obtained from the pathological archive of the University Hospital Regensburg and contained 393 pathological sections from 107 patients from the period 2016 to 2018. The second cohort also originated from the pathological archive of the University Hospital Regensburg and contained 356 pathological sections from 95 patients from the period 2019 to 2021. The third cohort was obtained from the pathological archive of the University Medical Center Hamburg-Eppendorf. This cohort contained 189 pathological sections from 86 patients from the period 2019 to 2021. The fourth cohort was obtained from the pathological archive of the University Hospital Aachen containing 141 pathological sections from 37 patients from the period 1999 to 2014. Cohorts were consecutive retrospective case series. We pragmatically aimed to maximize the sample size of training and testing cohorts. The ground truth was obtained by two expert pathologists during routine work-up at each participating center, grading the degree of rejection in consensus, following the 2004 revision of the ISHLT grading system.[5] All patient samples without information on ISHLT grading were not eligible for inclusion. A detailed presentation of the clinical characteristics of all patients in the corresponding cohorts can be found in **Table 1**. We used two categorizations of the ISHLT 2004 grading system as our prediction target. The first is a binarized target (“ISHLT 2004 rejection “yes/no”), summarizing slides with ISHLT 0R on the one hand (class “no”) and all signs of rejection on the other hand (ISHLT 1R, 2R, 3R; class “yes”). For the second target (“ISHLT 2004 rejection grade”) we aimed for a more granular classification splitting the second class giving three classes comprising ISHLT 0R, ISHLT 1R and ISHLT 2R & 3R. We combined the higher order rejection due to shortage of ISHLT 3R cases in the training set (**Table 1**). Our study adheres to the STARD guidelines (**Suppl. Table 1**). [19]

**Table 1:** Clinical characteristics of all cohorts. N/A Not available

|  | **Cohort 1** | **Cohort 2** | **Cohort 3** | **Cohort 4** |
| --- | --- | --- | --- | --- |
| **Contributing centre** | Regensburg | Regensburg | Hamburg | Aachen |
| **Use in this study** | Train | Test | Test | Test |
| **N patients** | 107 | 95 | 86 | 37 |
| **N slides** | 393 | 356 | 189 | 141 |
| **Recruitment years** | 2016 - 2018 | 2019 - 2021 | 2019 - 2021 | 1999 - 2014 |
| **Age in years (median, IQR)** | N/A | 61 (10) | 52 (17) | 55 (12) |
| **Gender (F:M)** | N/A | N/A | 70:121 | 47:94 |
| **ISHLT rejection** |  |  |  |  |
| no | 312 | 271 | 130 | 84 |
| yes | 81 | 85 | 59 | 57 |
| **ISHLT 0R** | 312 | 271 | 130 | 57 |
| **ISHLT 1R** | 51 | 77 | 51 | 24 |
| **ISHLT 2/3R** | 30 | 8 | 8 | 60 |

### Sample processing and image preprocessing

Routine tissue sections were obtained from the pathology archives at the above-mentioned institutions. All slides were stained with hematoxylin and eosin (H&E) according to standard clinical protocols at each center. Pen marks were removed from the slides of the training cohort. All images were digitized at 40x magnification with an Aperio AT2 Slide scanner (Aperio, Leica Camera AG, Wetzlar, Germany) centrally at the University Hospital Düsseldorf (**Figure 1a**). All images were available in ScanScope Virtual Slide (SVS) format and were tessellated in tissue patches of 512×512 pixels size using https://github.com/KatherLab/preprocessing-ng according to the “The Aachen Protocol for Deep Learning Histopathology: A hands-on guide for data preprocessing” (**Figure 1b**) [20].

**Figure 1:**
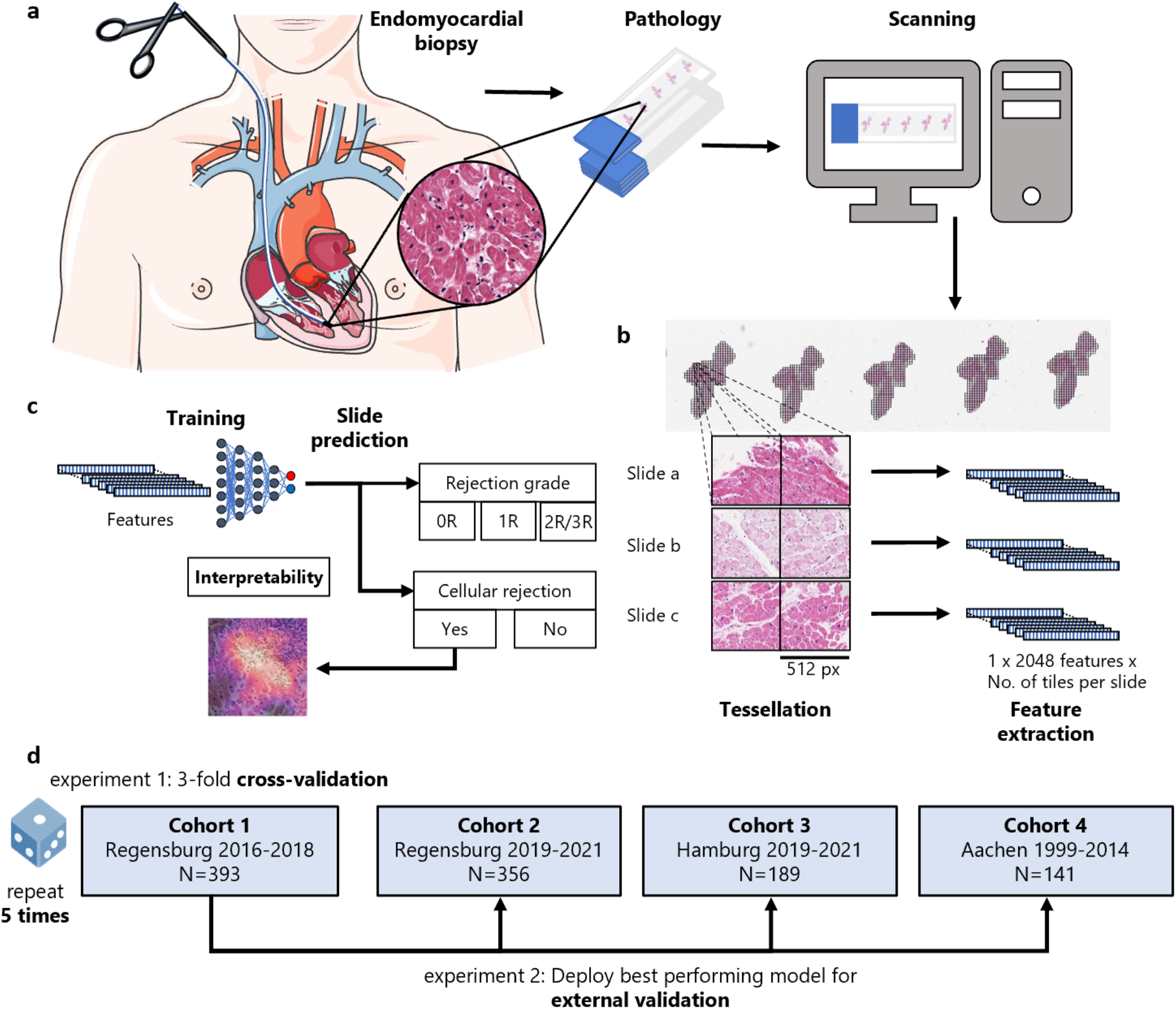
Outline of the study procedures. **a)** Routine endomyocardial biopsies of the right interventricular septum were taken from heart transplanted patients. These biopsies were then prepared into H&E stained histopathology slides, before being digitized and turned into whole slide images (WSIs) by use of a slide scanner. (Icon from smart.servier.com) **b)** To make these WSIs processable for our attention-based deep learning models, in a first step they need to be cut into smaller tiles while the background and artifacts are removed (tessellation). In the next step feature maps are extracted from all tiles from all slides using a publicly available neural network, which has been pre-trained by self-supervised learning with thousands of histopathology images. **c)** The resulting bags of feature maps per slide, together with expert pathologists’ opinion on the occurrence of rejection on a slide level as target label, are then used as training input for an attention-based deep learning model. **d)** In a first experiment, three-fold cross-validation is performed within Cohort 1 and repeated 5 times. In a second experiment the best performing model from experiment 1 is externally validated on Cohorts 2, 3 and 4.

### Deep Learning workflow

For all Deep Learning experiments, we used our in-house pipeline “Marugoto”, which is publicly available at https://github.com/KatherLab/marugoto and has been previously used for analysis of images obtained from cancer tissue [21]. In this approach, each image tile was translated into a 2048-dimensional feature vector by a pre-trained histology-specific encoder RetCCL (https://github.com/Xiyue-Wang/RetCCL). We used attention-based multiple instance learning [22], in which all feature vectors obtained from all tiles from one whole slide image constitute a “bag” which is processed by the neural network (**Figure 1b**). The multiple instance learning network is structured as follows: The feature vectors of each of the bag’s tiles are first projected into a length 256 feature space using a fully connected layer. Based on these, an attention module consisting of two fully connected layers calculates an attention score for each of the tiles. All of a bag’s attention scores are then normalized using softmax. We then calculate a bag-level feature vector by taking the sum of the tiles’ feature projections weighted by their respective attention scores. The final classification is then done with an additional fully connected layer (**Figure 1c**). During training, we limited our bag size to a maximum of 512 tiles from each slide, resampled in each epoch (median number of tiles = 403, interquartile range = 468). We stopped the training of our model if no reduction in the validation loss was present for 16 following epochs while training for a maximum of 32 epochs. For deployment, we used all of the slides’ tiles. We compared our approach to CRANE as presented by Lipkova et al. [16]. To do so, we followed the workflow of the CRANE study, preprocessing the slides with the CLAM repository and performed 10-fold Monte-Carlo cross validation on our training cohort, deploying the best performing model on our test cohorts [23].

### Experimental design and hardware

We pre-specified the following experimental design. First we trained and evaluated our system in the first cohort via three-fold cross validation and repeated this experiment five times. Subsequently, we evaluated the performance of the best performing model on the second, third and fourth cohort (**Figure 1d**). All ground truth labels were available on the level of slides. All statistics were calculated on the level of slides. The primary evaluation metric was the Area Under the Receiver Operating Curve (AUROC). We calculated the mean performance as the mean of all AUROCs from all folds of all repetitions, together with the 95% confidence interval (95% CI) calculated by assuming a normalized distribution of AUROCs and using its standard error of the mean to identify the boundaries. For the multiclass prediction we used micro-averaging to obtain an overall AUROC of the experiments. We calculated p-values for each class in each experiment using a two sided t-test and averaged these values over folds and repetitions of the experiments. For visualization approaches, we deployed the best performing model on the test cohorts. All experiments were run on local desktop workstations with Nvidia RTX Quadro 8000 graphics processing units (GPUs).

### Visualization and explainability

We plotted three tiles for the four slides of each validation cohort giving the highest bag label scores for the binarized prediction of (true) rejection when deploying the best performing model. Additionally we generated Grad-CAM images for these tiles to get a better understanding of the models attention.[24] To gain further insight into our model’s decisions, we generated heatmaps showing the attention, as well as the attention multiplied by the prediction scores.

### Code availability

All source codes for preprocessing are available at https://github.com/KatherLab/preprocessing-ng. All source codes for Deep Learning are available at https://github.com/KatherLab/marugoto.

## Results

### Deep learning can predict rejection and rejection grade from pathology images

We trained an attention-based multiple-instance deep learning algorithm on bags of features, extracted from patches of whole slide images. In the cross-validated experiment carried out on cohort 1, we found a mean AUROC of 0.849 (95% CI 0.822 - 0.877) for binary prediction (rejection yes/no) (**Figure 2a**, see **Suppl. Table 2** for individual results). The best fold’s AUROC was 0.910 with a p-value of <0.001. For prediction of the ISHLT grades 0R, 1R and 2/3R, the mean AUROCs were 0.835 (95% CI 0.807 - 0.862), 0.633 (95% CI 0.582 - 0.684) and 0.905 (95% CI 0.874 - 0.937), respectively (**Figure 2b**, see **Suppl. Table 3** for individual results). The micro-averaged AUROC for this task was 0.814 (95% CI 0.773 - 0.854). The best-fold’s AUROCs for this task were 0.890, 0.808 and 0.968, respectively, with a p-value <0.001 and a micro-averaged AUROC of 0.885. These results show the capacity of our network to predict rejection and rejection grade directly from histopathology images.

**Figure 2:**
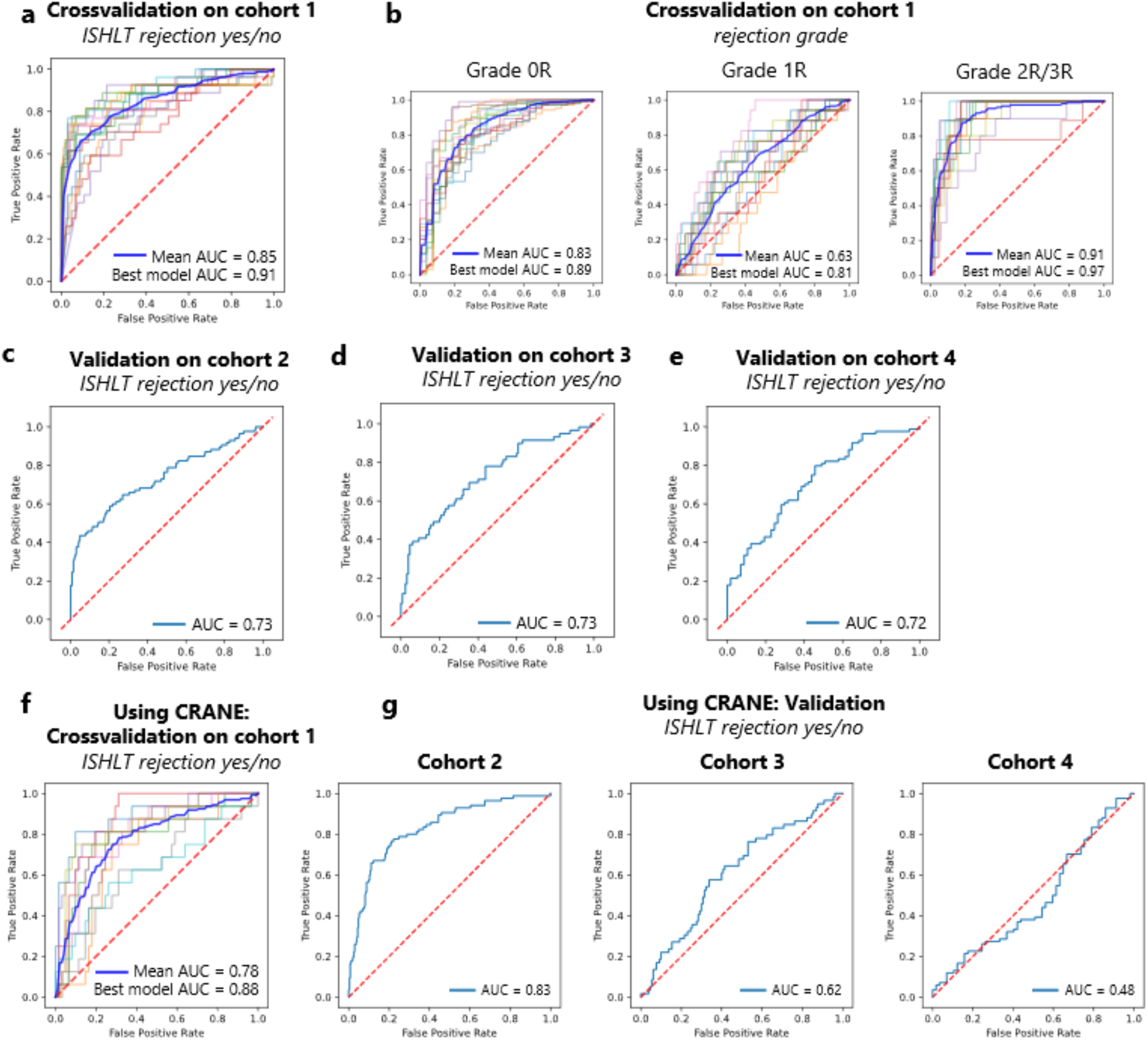
Deep learning can predict rejection and rejection grade from pathology images. Receiver operator characteristic curves (ROC) and mean area under the receiver operator curve. as measure of performance of the classifier for heart transplant rejection following 2004 revision of the International Society for Heart and Lung Transplantation (ISHLT) grading system. Showing binarized prediction (ISHLT rejection yes/no) (**a, c, d**, and **e**) and rejection grade (ISHLT 0R, 1R, 2/3R) (**b**) for cross-validation (**a** and **b**) and external validation (**c, d**, and **e**) experiments, as well as cross validation (**i**) and external validation (**j**) for binarized prediction (ISHLT rejection yes/no) using CRANE algorithm.

### Deep learning classifiers generalize to held-out and external patient cohorts

To further validate the performance of our network, we deployed the best performing model for each target on three validation cohorts. The validation experiments for cohort 2 yielded an AUROC of 0.734 (p-value <0.001) for binary prediction (rejection yes/no) (**Figure 2c**). For prediction of the ISHLT grades 0R, 1R and 2/3R, the AUROCs were 0.764 (p-value <0.001), 0.597 (p-value 0.099) and 0.913 (p-value <0.001) (**Suppl. Figure 1a**), respectively. The micro-averaged AUROC was 0.731 (p-value 0.021). For external validation on cohort 3 we obtained an AUROC of 0.729 (p-value of <0.001) (**Figure 2d**). For prediction of the ISHLT grades 0R, 1R and 2/3R, the AUROCs were 0.631 (p-value <0.013), 0.595 (p-value 0.048) and 0.682 (p-value 0.082), respectively (**Suppl. Figure 1b**). The micro-averaged AUROC was 0.659 (p-value 0.025). The external validation on cohort 4 yielded an AUROC of 0.716 (p-value <0.001) on the binary task (rejection yes/no) (**Figure 2e**). For prediction of the ISHLT grades 0R, 1R and 2/3R, the AUROCs were 0.722 (p-value <0.001), 0.601 (p-value 0.247) and 0.805 (p-value <0.001), respectively (**Suppl. Figure 1c**). The micro-averaged AUROC was 0.737 (p-value 0.042). Our findings show that our models are in principle generalizable to external patient cohorts.

### Comparison of the deep learning classifier with CRANE

We compared our method to CRANE, the current state of the art in rejection prediction of heart transplant tissue slides [16]. In the training cohort, the cross-validated mean AUROC of the CRANE models for the binarized target (rejection yes/no) was 0.776 (95% CI 0.717 - 0.835) (**Figure 2f**, see **Suppl. Table 4** for individual results), lower than the performance obtained by our attention-MIL pipeline (0.849). The best performing CRANE model yielded an AUROC of 0.882, which was again slightly lower than the performance achieved by our in-house attention-MIL pipeline (0.910). When deploying the CRANE model to our test cohorts we received AUROCs of 0.831, 0.616, and 0.483 for cohorts 2, 3 and 4, respectively (**Figure 2g**), overall underperforming compared to our SSL-attention model (which reached 0.734, 0.729 and 0.716, respectively). In summary, our findings show that SSL-attention-MIL outperforms CRANE.

### Attention-based predictions are explainable

To make the model’s prediction explainable and to identify reasons for failure cases, we performed a reverse engineering task to see the spatial distribution of the network’s attention layer for the most confident true classification of binary prediction. First of all our attention maps show that our model is concentrating only on tissue regions and not on the background or artifacts (See **Figure 3c** and **3g**). This means that the presence of such artifacts (e.g., pen marks) in the test set is not problematic, and that only a simple quality control algorithm might be sufficient for clinical implementation. Analyzing whole slide attention and prediction maps on a higher resolution, we found that our model’s focus lies mainly on regions with a high lymphocyte density. Yet it seems to focus more on the interface of lymphocyte aggregations with the neighboring myocardium than on these dense regions themselves (see **Figure 3**). We also found evidence that our model apparently was confused by the presence of a Quilty lesion [25], which was observed in a misclassified patient (**Suppl. Figure 2**).

**Figure 3:**
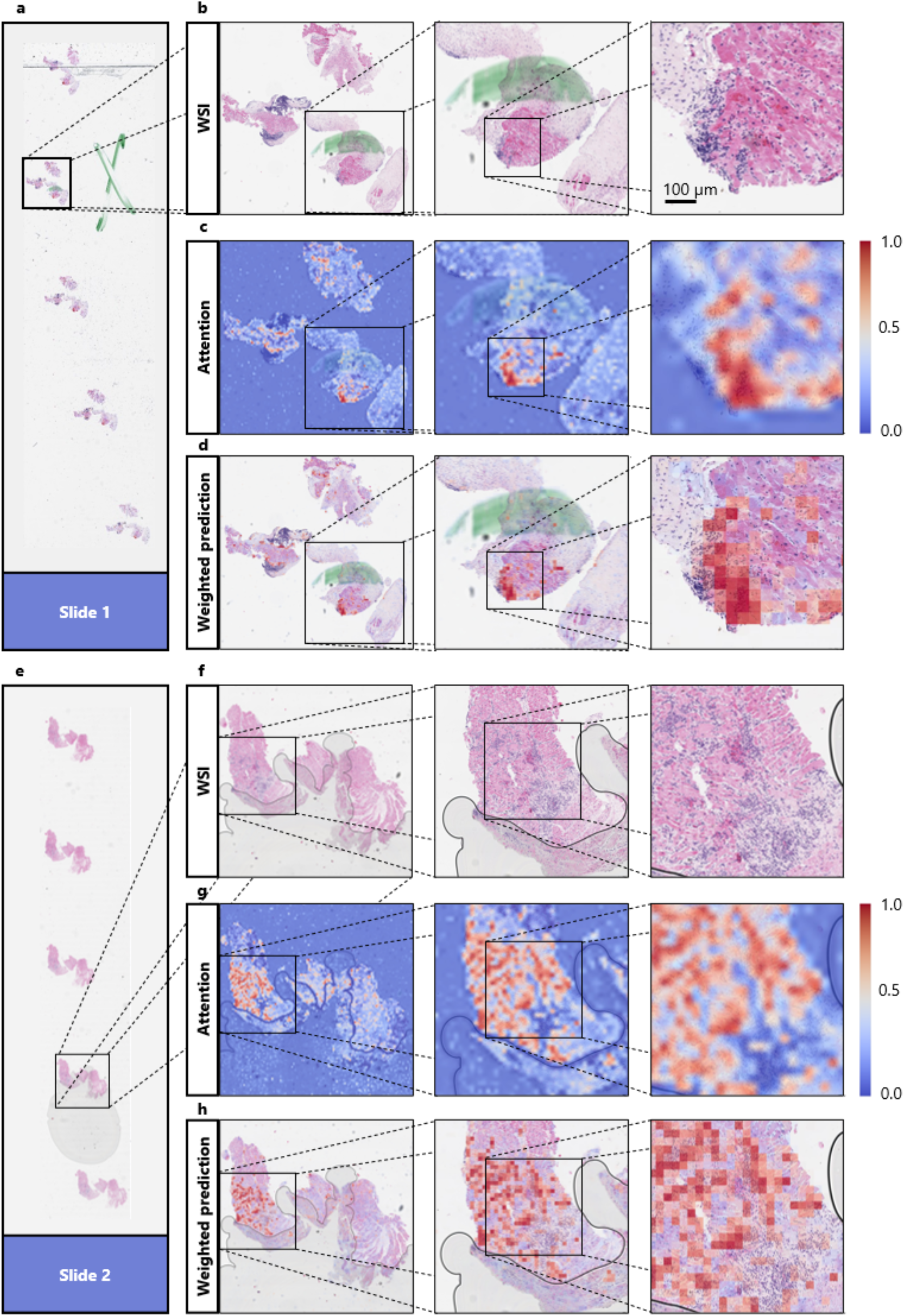
Explaining the models’ decisions by visualizing the model’s high attention regions. Different zoom levels of areas of the whole-slide-images (**b** and **f**) containing one patch of the endomyocardial biopsy of two different slides (**a** and **e**) together with the attention-based heatmap of the corresponding slide region (**c** and **g**) and a heatmap showing the attention scores multiplied by the prediction scores (**d** and **h**). In attention-based heatmaps dark red indicates regions with a high attention, while dark blue indicates regions with a low attention [see scale in **c)** and **g**]. The network is focussing on areas of the whole slide image containing tissue, ignoring artefacts, like air bubbles and pen marks (**d** and **h**). The network was trained on cohort 1 for the binarized target (rejection yes/no) and deployed on cohort 2. For those two slides, the network was the “the most confident” about its decision (reflected by highest attention and prediction scores). The network is highlighting regions with a high number of lymphocytes between heart muscle tissue.

When analyzing the top tiles and the corresponding Grad-CAM images of the external validation cohorts, it seems that the model is concentrating on lymphocytes, confirming the findings made in heatmaps at another spatial scale (**Figure 4**). These findings show that despite being trained on only a few hundred patients, the model has learned clinically relevant morphological patterns from whole slide images.

**Figure 4:**
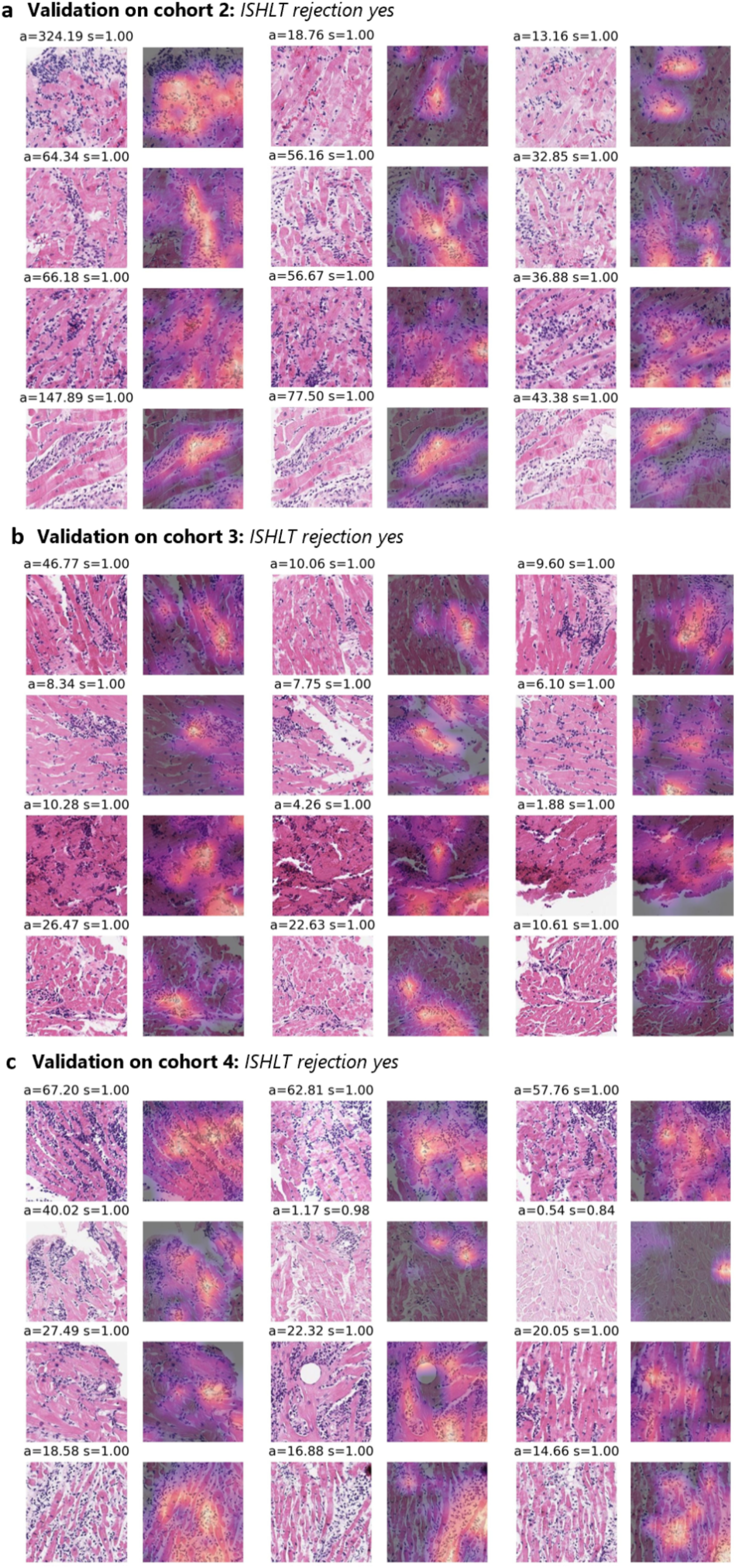
Explaining the models’ decisions by visualizing the model’s high attention 512×512 tiles. The three tiles (columns) with the highest average attention and prediction scores (attention = a, prediction score = s) for the four slides(rows) with the highest average prediction scores when deploying the best performing model to detect rejection (rejection yes/no) on the three test cohorts (**a**,**b**, and **c**). Together with the corresponding Grad-CAM images showing the network’s spatial attention for each of the tiles. Regions with higher attention are yellow, while regions with low attention are in dark purple. The top tiles contain many lymphocytes infiltrating the myocytes, while the network’s attention also appears to be lying on these immune cells.

## Discussion

Heart transplantation remains the gold standard therapy for end-stage heart failure [26]. Due to this pronounced shortage of donor organs, there is not only a need for risk adjustment tools to optimize recipient selection [27,28]. In addition, a particularly good risk stratification and early adjustment of immunosuppression therapy is necessary in organ recipients because the possibility of re-transplantation is very limited. New diagnostic methods based on artificial intelligence (AI) could change and improve medical decision making in transplantation medicine in the future.[17] A potential key benefit would be to reduce diagnostic uncertainty, and hence reduce the need for frequent re-biopsies in the first year after transplantation, which represents a burden for healthcare systems and patients alike.

In the present study, we trained an AI method to evaluate the recognition and grading of cardiac transplantation using routine biopsies. We found high performance in the training set (by cross-validation). When deploying our model at the external validation cohorts, we found a stable, but moderate performance. A few other studies have addressed similar problems in recent years. Peyster et al. used handcrafted features to grade cellular rejection reporting good performance, already in 2021 [15]. Most prominently, Lipkova et al. presented the CRANE method, which yielded very high AUROCs in their study, after being trained on thousands of patients [16]. Lipkova et al. report an external validation AUROC of around 0.83, which is better than the AUROCs of around 0.72 which we report in the validation cohorts [16]. However, our training dataset comprised 10 times fewer patients, and in a head-to-head comparison of CRANE and our SSL-attention, our method outperforms CRANE, pointing to a higher data efficiency. Our findings are in line with other recent studies showing the usefulness of pre-training feature extractors with SSL, boosting classification performance in computational pathology [18]. Our classifier also outperforms other studies which date back to the year 2017, when Tong et al. constructed a shallow neural network based on handcrafted features derived from 43 WSIs (Children’s Healthcare of Atlanta cohort). This dataset has been used several times afterwards improving the performance of the cross validated model while adopting newer methodology but remains limited due to the very small dataset size [14,29–31].

A fundamental limitation affecting all published studies is the limitation of the gold standard. The ISHLT classification itself is an imperfect predictor of clinical outcome, and future studies should train AI models directly on outcome data to overcome these limitations. This is further supported by the observation that detection and grading of heart transplant rejection can suffer from a suboptimal concordance among pathologists in the assignment of ISHLT 2004 grading of 71%, with most agreement coming from the class 0R [32]. While our study does not directly show that AI can improve objectivity and concordance, future studies should investigate the performance of pathologists who are guided by the AI model, especially non-expert pathologists.

In summary, our study is a proof of concept that shows the potential of AI systems in transplantation medicine. In particular, our study sets a new technical state of the art, which however requires validation in larger cohorts. On the other hand, our study is also a reminder that larger training cohorts of a few thousand patients are probably required for clinical-grade AI biomarkers [33,34].

Future studies should compare our technical approach on larger cohorts, which could be efficiently assembled with federated or Swarm Learning [35,36]. Our study adds to the growing evidence of AI models being capable of recognizing heart transplant rejection which might in the future help pathologists with prescreening slides or standardize grading across different centers. We also believe that further development of our approach harbors the potential to ultimately reduce costs and time in this sector of the healthcare system.

## Data Availability

All data produced in the present work are contained in the manuscript.

## Additional information

### Author contributions

TPS, ML, CR, CB, SS and JNK conceived the idea for the study. TPS and JNK performed the experiments. MVT developed the software. PS, RDB, PB, ZP, DM, FB, CM, DW, CB and SS contributed tissue samples. TPS and JNK wrote the initial draft of the manuscript. All authors contributed to the interpretation of the results, the editing of the final manuscript and gave approval for submission.

## Declarations

### Ethics approval

This study was carried out in accordance with the Declaration of Helsinki. This study is a retrospective analysis of digital images of anonymized archival tissue samples from three patient cohorts. Collection and anonymization of patients in all cohorts took place in each contributing centre. For the Regensburg cohorts, ethical approval was granted by the ethical Review board of the University Regensburg (ID: 21-2620-104). For the Hamburg and Aachen cohort, ethical approval of this retrospective investigation was not legally required due to local regulations (Berufsordnung fuer Aerzte). Nevertheless, the retrospective analysis of samples from Aachen and other collaborating centers was assessed by and approved by the Ethics commission of the Medical Faculty of RWTH Aachen University (EK315/19), confirming that no specific patient consent is required for this retrospective study of anonymized tissue samples.

