## Supplementary material for "Prediction of heart transplant rejection from routine pathology slides with self-supervised Deep Learning": Suppl. Table Legends

### Supplementary Table Legends

[Suppl-Table-1.docx] in separate file

**Supplementary Table 1: STARD checklist.**

[Suppl-Table-2.xlsx] in separate file

**Supplementary Table 2:** **Results of the best performing model of each fold for each run in the cross-validation experiment on cohort 1 predicting binarized ISHLT 2004 rejection (rejection yes/no).**

[Suppl-Table-3.xlsx] in separate file

**Supplementary Table 3: Results of the best performing model of each fold for each run in the cross-validation experiment on cohort 1 predicting ISHLT 2004 rejection grade (0R, 1R, 2R/3R).**

[Suppl-Table-4.xlsx] in separate file

**Supplementary Table 4: Results of the best performing model of each fold for each run in the cross-validation experiment on cohort 1 using CRANE algorithm predicting binarized ISHLT 2004 rejection (rejection yes/no).**
