## Supplementary material for "Prediction of heart transplant rejection from routine pathology slides with self-supervised Deep Learning": Suppl. Figures

### Supplementary Figures

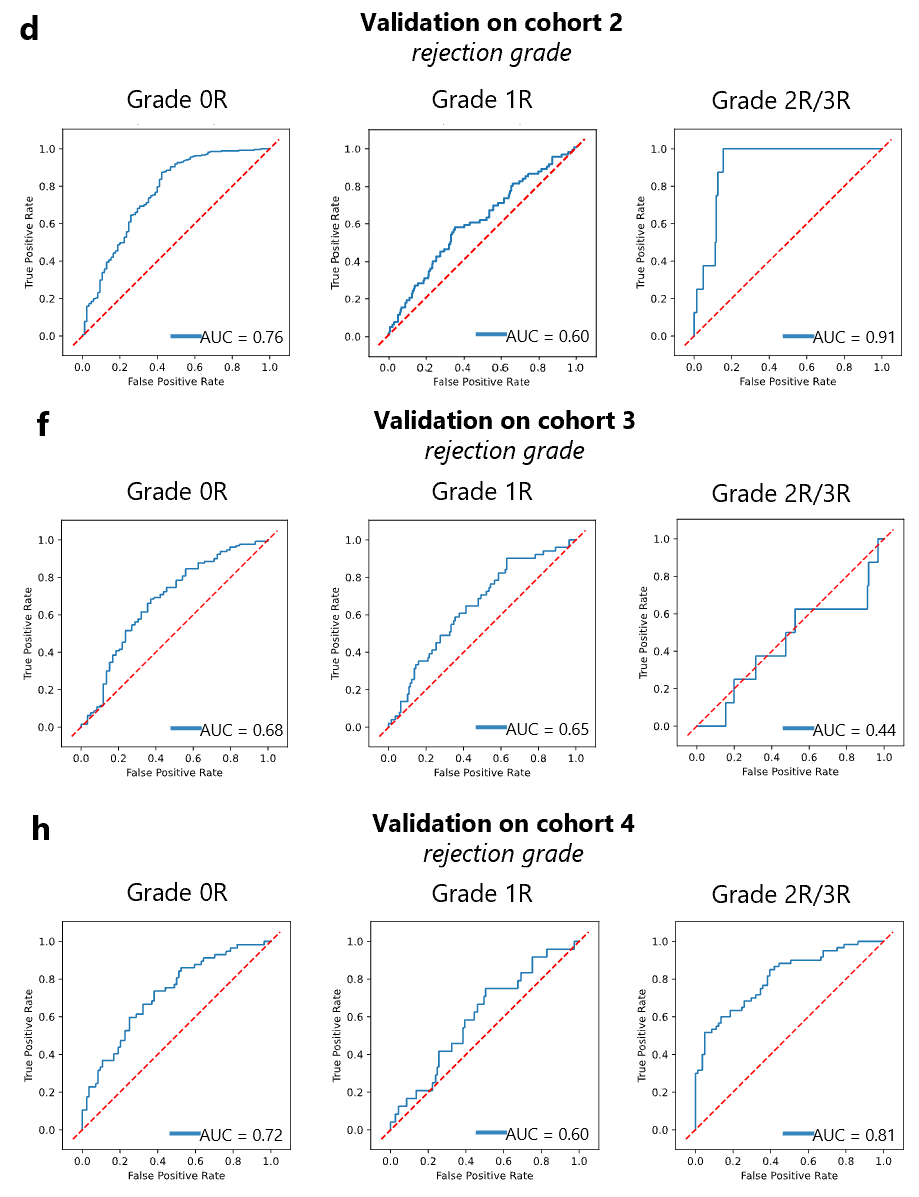

**Supplement Figure 1: Deep learning can predict rejection grade from pathology images.** Receiver operator characteristic curves (ROC) and area under the receiver operator curve (AUC), as measure of performance of the classifier for heart transplant rejection following 2004 revision of the International Society for Heart and Lung Transplantation (ISHLT) grading system. Showing rejection grade (ISHLT 0R, 1R, 2/3R) for external validation in cohorts 2,3 and 4 (**a**, **b**, and **c**).

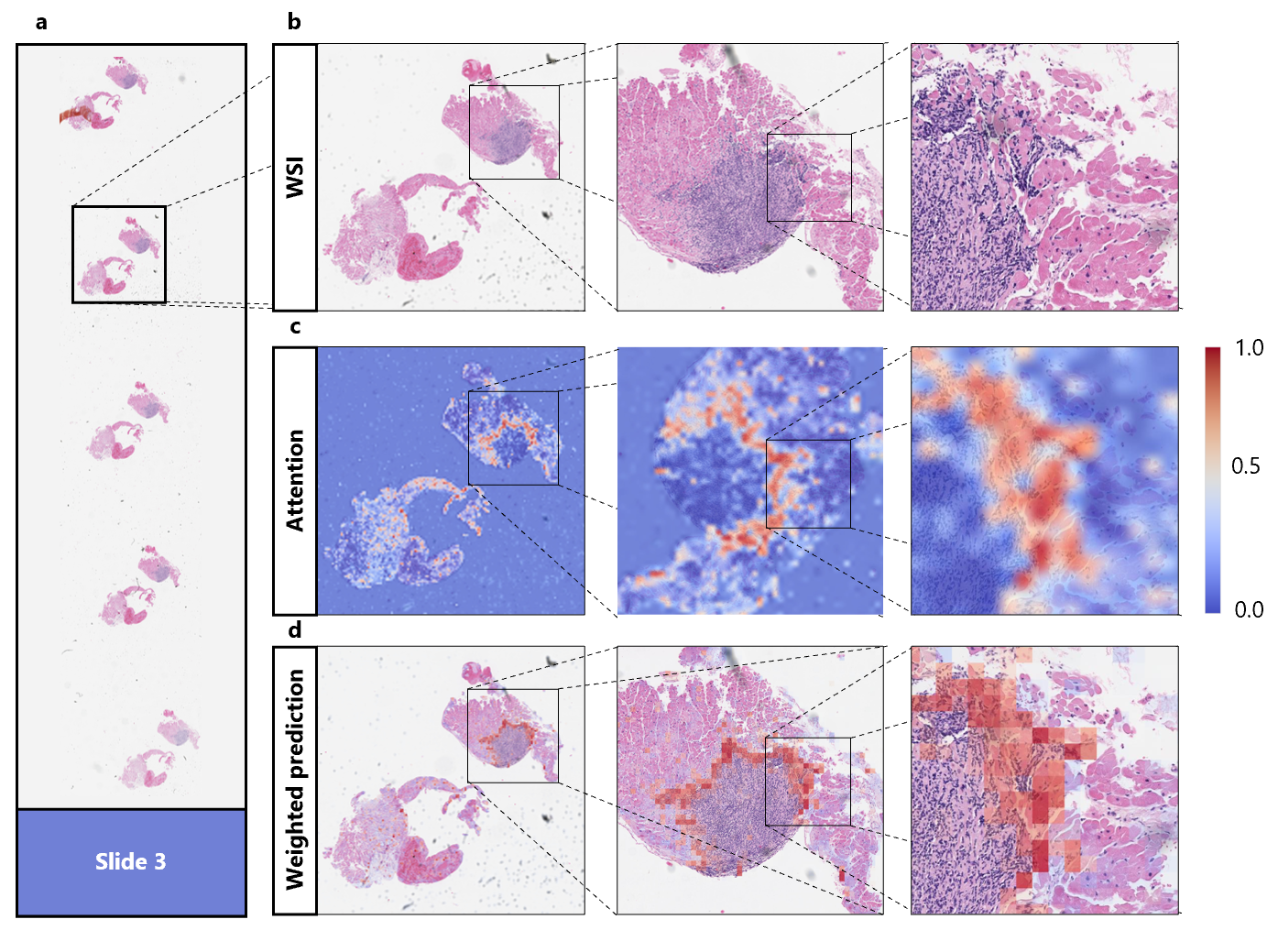

### **Supplement Figure 2: Explaining the models' decisions by visualizing the model’s high attention regions.** Different zoom levels of areas of the whole-slide-image (**b**) containing one patch of the endomyocardial biopsy the slide (**a**) together with the attention-based heatmap of the corresponding slide region (**c**) and a heatmap showing the attention scores multiplied by the prediction scores (**d**). In attention-based heatmaps dark red indicates regions with a high attention, while dark blue indicates regions with a low attention (see scale in **c**). The network was trained on cohort 1 for the binarized target (rejection yes/no) and deployed on cohort 2. This slide shows no signs of acute rejection, but a so-called Quilty lesion. Nevertheless it is classified as a rejection case by our classifier.
